# Comparing the Safety and Immunogenicity of homologous (Sputnik V) and heterologous (BNT162B2) COVID-19 prime-boost vaccination

**DOI:** 10.1101/2022.08.24.22279160

**Authors:** Marwa AlMadhi, Abdulla AlAwadhi, Nigel Stevenson, Khalid Greish, Jaleela Jawad, Adel Alsayyad, Afaf Mirza, Basma Alsaffar, Ejlal AlAlawi, Khulood Fakhroo, Batool Alalawi, Lana Alabbasi, Noora Aljalahma, Manaf AlQahtani

## Abstract

Studies have shown increased immunogenicity from heterologous boosting. This study specifically assessed boosting with Pfizer-BioNTech in Sputnik V vaccination regimens. Reactogenicity was assessed through adverse events. Immunogenicity was assessed by comparing serum anti-Spike (Anti-S) protein antibody and neutralizing antibody titers before booster administration and after 30 days. A total of 428 participants were recruited in the heterologous arm and 351 in the homologous arm. Adverse events were more frequent in the heterologous arm (p<0.001). No serious adverse events were reported in either arm. Amongst 577 individuals who completed the study, Anti-S antibodies were 14.8 times higher after heterologous boosting, and 3.1 times higher after homologous boosting (p<0.001). Similarly, heterologous boosting showed a 2 fold increase in neutralizing antibodies, compared to a 1.6 fold increase in homologous boosting (p<0.001). In conclusion, both boosting regimens elicited an immunological response, nonetheless heterologous Pfizer-BioNTech showed a higher immunological response, with more adverse effects.

**ARTICLE SUMMARY LINE:** Both homologous and heterologous boosting are effective in eliciting an immunological response, however heterologous boosting with Pfizer-BioNTech elicited a higher immunological response, with more adverse effects.

## INTRODUCTION

Severe Acute Respiratory Syndrome Coronavirus 2 (SARS-CoV-2) infection causes Coronavirus Disease 2019 (COVID-19). The outbreak emerged in China in December 2019 and was declared a pandemic by the World Health Organisation (WHO) in March 2020 (1). Over the past two years, COVID-19 has cumulated over 452,000,000 cases and 6,000,000 deaths worldwide (2). As restrictions and national lockdowns were enforced, researchers and pharmaceutical companies worldwide raced to discover effective and safe vaccines.

There are currently 148 COVID-19 vaccines in clinical trials and 195 in pre-clinical developments (3). Of the 10 vaccines granted approval by the WHO, over 10 billion vaccine doses have been administered as of 12 March 2022 (2). Based on their efficacy data, clinical trials have played a key role in the approval of these COVID-19 vaccines. However, despite the availability and uptake of these vaccines, the world has recently faced the largest wave of infection since the start of the pandemic, the B.1.1.529 (Omicron) wave. Indeed, Omicron reached over 4 million daily cases worldwide at its peak (2). Studies have shown that new SARS-CoV-2 variants reduce the efficacy of vaccinations and are predominantly more transmissible or infective (4). Reports of infections in fully vaccinated individuals began surfacing in parallel with the B.1.617.2 (Delta) variant, and continued through the latest Omicron wave (5, 6).

Recent preliminary results from a comparative study by Lapa D *et al*., reports an 8.1 fold drop in neutralizing antibodies in Sputnik V-vaccinated individuals, compared to a 21.4 fold decrease in Pfizer-BioNTech (BNT162b2) vaccinated individuals 3-6 months following vaccination (7). This study concludes a possibility that Sputnik V vaccine may be more immunogenic against COVID-19 than Pfizer-BioNTech vaccine. Both Sputnik V (viral vector) and Pfizer-BioNTech (mRNA vaccine) are among the currently approved COVID-19 vaccines in the Kingdom of Bahrain. Nonetheless, evidence for humoral response and immunogenicity following homologous vaccination is predominantly undisputed. However, the practical course of events during the pandemic prompted interest in heterologous vaccination. For instance, the rise in severe thrombotic events following AstraZeneca (ChAdOx1 nCoV-19) vaccines lead to some countries to halt its use and turn to heterologous boosting regimens. (8) Furthermore, national and global strains with maintaining vaccine supplies, coupled with the continuous emergence of new SARS-CoV-2 variants contributed to the growing interest in heterologous vaccinations. (9)

Propitiously, a relatively novel hypothesis suggests that responses may in reality be enhanced in heterologous vaccinations. In fact, several studies have shown heterologous vaccinations regimens to induce significantly more immunogenicity than that of homologous vector vaccinations, and be comparable or higher than homologous mRNA regimens. A study by *Liu et al*. compared homologous AstraZeneca vaccination with a heterologous vaccination regimen using Pfizer-BioNTech in a total of 830 individuals. (10). Immunogenicity was evaluated by quantitative comparison of antibodies titers to SARS-CoV-2 spike proteins, and neutralizing antibody titers. The study shows an approximate 9-fold increase in both antibody titers following heterologous vaccination compared to homologous AstraZeneca vaccination regimen. However, this was slightly lower than the immunogenicity reported for homologous Pfizer-BioNTech vaccination within the same study. (10) This effect was observed in several other studies (11, 12), however the immunogenicity of heterologous vaccination was consistently higher than in homologous vaccination of AstraZeneca. Other similar studies report the highest antibody titers amongst the groups with heterologous boosting compared to both homologous vaccination with AstraZeneca or Pfizer-BioNTech, with titers reported for homologous AstraZeneca vaccination regimens being the lowest. (13-15) A systemic review of 19 studies comparing heterologous vaccinated in COVID-19 showed that heterologous vaccination conferred more potent immunogenicity than homologous vaccination. (9) In addition, this was also observed against the delta variant.

Most studies have focused on heterologous boosting using AstraZeneca and Pfizer-BioNTech vaccines. In this study, we compared the reactogenic and immunogenic responses of homologous Sputnik V booster, versus heterologous Pfizer BioNTech booster following complete two-dose Sputnik V vaccination.

## METHODS

### 1. Study Design

This phase II non-randomized, non-blinded observational community trial was conducted across the Kingdom of Bahrain, investigating the reactogenic and immunogenetic response of heterologous versus homologous COVID-19 vaccine boosting of participants who previously received two doses of Sputnik V vaccine. The trail was 30 days in duration, conducted between September 14 2021 and October 14 2021. Participants received either a homologous Sputnik V C1 (Sputnik V) booster or BNT162b2 (Pfizer-BioNTech) heterologous booster on day 0, based on participant preference, and were followed up for 30 days.

The Pfizer-BioNTech vaccine used in this study has been available in Bahrain after an emergency use authorization was granted by the National Health Regulatory Authority (NHRA) in December 2020. The Pfizer-BioNTech booster was administered at the approved dose of 0.3 mL, the Sputnik V booster was administered at 0.5mL, as a single intramuscular injection. The Sputnik V vaccine used in this study has been available in Bahrain after an emergency use authorization was granted in February 2021. Participants were observed on-site for at least 15 minutes after vaccination for safety monitoring.

Reactogenicity was investigated using reports of adverse events. These were recorded through follow-up calls on day 1, 5 and weekly thereafter, till the completion of the 30-day study period. Participants were also instructed to record adverse events through an electronic diary throughout the study period. Adverse events included local reactions (hardness, itch, pain, warmth, redness, swelling) or systemic reactions (chills, fatigue, fever, headache, joint pain, malaise, muscle ache, nausea, vomiting, diarrhea). The intensity of adverse events was graded by the healthcare worker conducting the review as: Grade 1 (mild), Grade 2 (moderate), Grade 3 (severe), and Grade 4 (life-threatening), based on standardized description set by the primary investigator for each. Participants who missed the final follow-up call were called again on day 35.

Immunogenicity was investigated by measuring (i) Spike (S) antigen-specific humoral response, and (ii) sVNT pseudovirus neutralization from blood samples collected on day 0 (before booster administration) and day 30, following completion of the study period. The S antigen-specific (Anti-S) humoral immune response was analyzed using the Elecsys® Anti-SARS-CoV-2 S assay (Roche Diagnostics GmbH, Mannheim, Germany), which is an electrochemiluminescence immunoassay (ECLIA) that detects IgG antibodies to the SARS-CoV-2 spike protein receptor-binding domain (RBD) on the Cobas e411 module. According to the manufacturer, the measuring range spanned from 0.4 U/mL to 25,000 U/ml (up to 250 U/ml with on-board 1:10 dilution and more than 2,500 with onboard 1:100 dilution). Values higher than 0.8 BAU/mL were considered positive. The correlation between U/ml and BAU (International OMS standard) is U=0.972 BAU. sVNT psuedovirus neutralisation was analysed using cPass™ SARS-CoV-2 Neutralization Antibody Detection Kit (Genscript Biotech Corporation, Nanjing, China); neutralizing antibodies were calculated as 30% inhibitory dose (neutralizing titre 30, NT30). Neutralising antibodies were only measured in participants with anti-S titres >30 U/ml.

The study period ended 30 days after booster administration, where participants presented for blood sample collection and a final adverse event report. Patients who failed to attend the final review on day 30 were contacted again for review on day 35. The study timeline is summarized in the figure below

**Figure 1.**
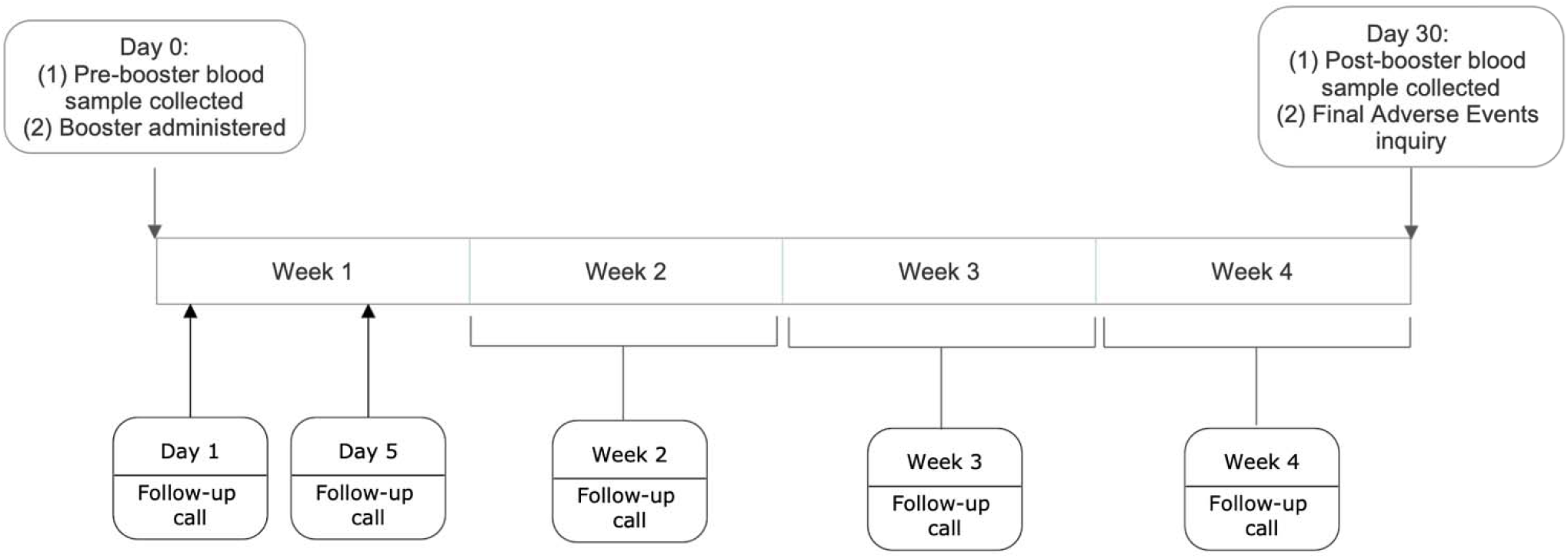
Summary of trial timeline. Events indicated by boxes above the timeline represent in-person consultations and appointments, while those below the timeline represent phone consultations. Individuals who did not attend the Day 30 appointment were offered further follow-up on Day 35.

### 2. Participants

First, using the Bahrain National COVID-19 database, we identified individuals in the Kingdom of Bahrain who received 2 doses of Sputnik V vaccine ≥ 6 months from the start of trial, and who were eligible for the booster vaccination dose. From this cohort, individuals who were aged ≥ 18 years, had no active or previous RT-PCR lab-confirmed COVID-19 diagnosis in 2021 were invited to participate. Of all eligible individuals identified, 779 accepted to receive the booster dose of the vaccine and participate in the trial. The 202 participants who withdrew before the end of the 30 day observation period were offered a follow-up within 4 weeks, to review adverse events.

Sample size was estimated using GPower version 3.1.9.7 (16). The minimum sample size estimated to reliably study heterologous boosting was 120 participants in each study arm at 80% power and an alpha of 0.05.

### 3. Outcomes

Primary outcome: reactogenicity of both heterologous and homologous boosting through assessment of adverse events.

Secondary outcomes: immunogenicity of heterologous boosting compared to homologous boosting through quantitative comparison of serology tested antibodies (Anti-S and neutralising antibodies) 30 days after booster administration.

### 4. Data Handling and Statistical Analysis

Epidemiological data is presented as n and %, where n is the sample size corresponding to each category, and % is the category sample size as a proportion of the total cohort studied, unless otherwise stated. Z-test of proportion was used to statistically compare two proportions. Chi-squared tests were used to compare more than two values, in contingency tables of 2×2 or more. P-values were considered statistically significant at p<0.05. SPSS version 27 was used for statistical analysis (IBM Corp. Released 2020. IBM SPSS Statistics for Windows, Version 27.0. Armonk, NY: IBM Corp). Sample size was estimated using GPower version 3.1.9.7

### 5. Ethical approval

The protocol and manuscript for this study were reviewed and approved by the National COVID-19 Research Committee in Bahrain (CRT-COVID2021-153). All methods and data analysis were approved by the National COVID-19 Research and Ethics Committee. The study was carried out following local guidelines and in line with the 2013 Declaration of Helsinki.

## RESULTS

### 1. Primary analysis of cohort

A total of 779 individuals were included in the study, of which the majority were male (n=549, 70.5%), less than 40 years old (n=443, 56.9%). Of the 779 participants, 428 (54.9%) chose the heterologous Pfizer-BioNTech arm and 351 (45.1%) chose the homologous Sputnik V arm, of the booster trial. Most participants were male and less than 40 years old in both arms of the trial.

### 2. Safety: Reactogenicity

We assessed reactogenicity through incidence of adverse events. We observed a statistically significant difference in the number of adverse events reported between heterologous and homologous boosting (p<0.001), with a total of 161 (37.6%) participants reporting adverse events to the Pfizer-BioNTech booster, and 76 (21.7%) to the Sputnik V booster.

Local adverse events at the injection site were the most commonly reported in participants with heterologous boosting (n=79, 18.5%). This was also the adverse event with highest incidence for both <40 years (20.1%) and 40-59 years age groups (17.3%). No adverse events were reported with heterologous boosting in participants aged 60 and above. The lowest incidence of adverse event was fever (n=38, 8.9%), which also was the lowest in incidence for both <40 years (9.2%) and 40-59 years (8.9%) age groups. For homologous boosting, local adverse events and systemic, non-injection site adverse events were both of equal incidence (n=31, 8.8%). Local adverse events were of highest incidence in the <40 year age group (12.4%), while systemic adverse events were the highest reported in the 40-59 age group (7.7%). Only 1 adverse event (6.7%), was reported in participants aged 60 and above, in homologous boosting. Fever was the adverse event with lowest incidence overall (n=11, 3.1%), and in participants aged 40-59 (1.4%). The lowest incidence of adverse events in participants aged <40 corresponded to the “other symptoms” category (3.6%). No serious adverse events were reported in this study. Adverse events reported for each arm of the booster trial are summarised in Table 2.

**Table 1.** Summary of trial participant demographics*.

|  |  | **Total**<br>**n (%)** | **Heterologous**<br>**(Pfizer-BioNTech)**<br>**n (%)** | **Homologous**<br>**(Sputnik V)**<br>**n (%)** | **P-values** |
| --- | --- | --- | --- | --- | --- |
| Gender | Male | 549 (70.5) | 297 (69.4) | 252 (71.8) | 0.5 |
|  | Female | 230 (29.5) | 131 (30.6) | 99 (28.2) |  |
| Age<br>group<br>(years) | < 40 | 443 (56.9) | 249 (58.2) | 194 (55.3) | 0.36 |
|  | 40 – 59 | 103 (39.8) | 168 (39.3) | 142 (40.5) |  |
|  | 60+ | 26 (3.3) | 11 (2.6) | 15 (4.3) |  |
| **Total** |  | **779** | **428** | **351** |  |

\* Demographics are reported as total, and for each arm of the trial. Values are expressed as the n, (%), where n represents the number of participants and % represents the corresponding number of participants as a proportion of the column total

**Table 2.** Summary of adverse events reported in heterologous and homologous boosting.

|  | N | No. of participants | No. of events | Incidence, % | N | No. of participants | No. of events | Incidence, % | p-value |
| --- | --- | --- | --- | --- | --- | --- | --- | --- | --- |
| **Fever** | **428** | **38** | **38** | **8.9** | **351** | **11** | **14** | **3.1** | **0.001** |
| Age group: |  |  |  |  |  |  |  |  |  |
| <40 | 249 | 23 | 23 | 9.2 | 194 | 9 | 11 | 4.6 |  |
| 40 – 59 | 168 | 15 | 15 | 8.9 | 142 | 2 | 3 | 1.4 |  |
| 60+ | 11 | 0 | 0 | 0.0 | 15 | 0 | 0 | 0.0 |  |
| **Injection Site (Local)** | **428** | **79** | **83** | **18.5** | **351** | **31** | **31** | **8.8** | **<0.001** |
| Age group: |  |  |  |  |  |  |  |  |  |
| <40 | 249 | 50 | 51 | 20.1 | 194 | 24 | 24 | 12.4 |  |
| 40-59 | 168 | 29 | 32 | 17.3 | 142 | 6 | 6 | 4.2 |  |
| 60+ | 11 | 0 | 0 | 0.0 | 15 | 1 | 1 | 6.7 |  |
| **Non-injection Site (Systemic)** | **428** | **57** | **66** | **13.3** | **351** | **31** | **31** | **8.8** | **0.049** |
| Age group: |  |  |  |  |  |  |  |  |  |
| <40 | 249 | 33 | 38 | 13.3 | 194 | 20 | 20 | 10.3 |  |
| 40-59 | 168 | 24 | 28 | 14.3 | 142 | 11 | 11 | 7.7 |  |
| 60+ | 11 | 0 | 0 | 0.0 | 15 | 0 | 0 | 0.0 |  |
| **Other symptoms** | **428** | **42** | **48** | **9.8** | **351** | **12** | **13** | **3.4** | **<0.001** |
| Age group: |  |  |  |  |  |  |  |  |  |
| <40 | 249 | 26 | 30 | 10.4 | 194 | 7 | 8 | 3.6 |  |
| 40-59 | 168 | 16 | 18 | 9.5 | 142 | 5 | 5 | 3.5 |  |
| 60+ | 11 | 0 | 0 | 0.0 | 15 | 0 | 0 | 0.0 |  |
| **Overall** | **428** | **161** | **235** | **37.6** | **351** | **76** | **90** | **21.7** |  |

\*Incidence of each event was calculated using the corresponding number of participants as a proportion of the total participants in each category, indicated by the row total in the “n” column for each trial arm. p-values were calculated using paired sample t-test.

Next, we stratified adverse events by time of occurrence after booster administration (Table 3). The highest number of events were reported on day 1 for both heterologous (n=145) and homologous (n=54) boosting, with the majority being local injection site adverse events (n=70 and n=25, respectively). The least number of adverse events were reported on day 0 in both trial arms. After peaking at day 1, the number of adverse events reported generally decreased, before peaking again at day 28. This was observed in both heterologous and homologous boosting.

**Table 3.** Summary of number of adverse events reported in heterologous and homologous boosting by type of event and time from booster administration*.

|  | N | Heterologous<br>(Pfizer-BioNTech) |  |  |  |  |  |  | N | Homologous<br>(Sputnik V) |  |  |  |  |  |  |
| --- | --- | --- | --- | --- | --- | --- | --- | --- | --- | --- | --- | --- | --- | --- | --- | --- |
|  |  | Day 0 | Day 1 | Day 5 | Day 14 | Day 21 | Day 28 | Day 35 |  | Day 0 | Day 1 | Day 5 | Day 14 | Day 21 | Day 28 | Day 35 |
| **Fever** | **38** | - | 18 | 3 | 1 | - | 16 | - | **14** | - | 10 | - | - | 1 | 3 | - |
| **Injection Site**<br>**(Local)** | **83** | 1 | 70 | 4 | - | 1 | 7 | - | **31** | - | 25 | 1 | - | - | 5 | - |
| **Non-injection**<br>**Site**<br>**(Systemic)** | **66** | - | 36 | 11 | 2 | 4 | 13 | - | **31** | - | 18 | - | 5 | 2 | 5 | 1 |
| **Other**<br>**symptoms** | **48** | - | 21 | 5 | 3 | 5 | 12 | 2 | **13** | - | 1 | 4 | 2 | 1 | 5 | - |
| **Overall** | **235** | **1** | **145** | **23** | **6** | **10** | **48** | **2** | **89** | **0** | **54** | **5** | **7** | **4** | **18** | **1** |

\*'n' represents the number of events reported for each type of adverse event. Days with no adverse events reported are indicated by the '-'

### 3. Immunogenicity

We next assessed immunogenicity by comparing levels of Anti-S and neutralizing antibodies in heterologous and homologous boosted participants, before and 30 days after booster administration (Table 4). 202 participants withdrew from the trial before completion, leaving 319 participants in the heterologous Pfizer boosting cohort and 258 in the homologous Sputnik V boosting cohort. Of the remaining 577 participants, we observed an overall statistically significant 8.5-fold increase in Anti-S antibodies (p<0.001) and 1.8-fold increase in neutralizing antibody (p<0.001) from baseline. We observed a difference in the magnitude of antibody response when we analysed each trial arm separately (Table 4). Anti-S antibodies increased 14.8 fold in heterologous boosting, compared to 3.1 folds in homologous boosting, from baseline values (p<0.001). Baseline pre-booster levels of Anti-S antibodies were comparable between the two arms (p=0.052). Neutralizing antibodies increased 2 fold in heterologous boosting compared to 1.6 fold in homologous testing, from baseline (p<0.001). However, baseline pre-booster levels of neutralizing antibodies were significantly higher in

**Table 4.**
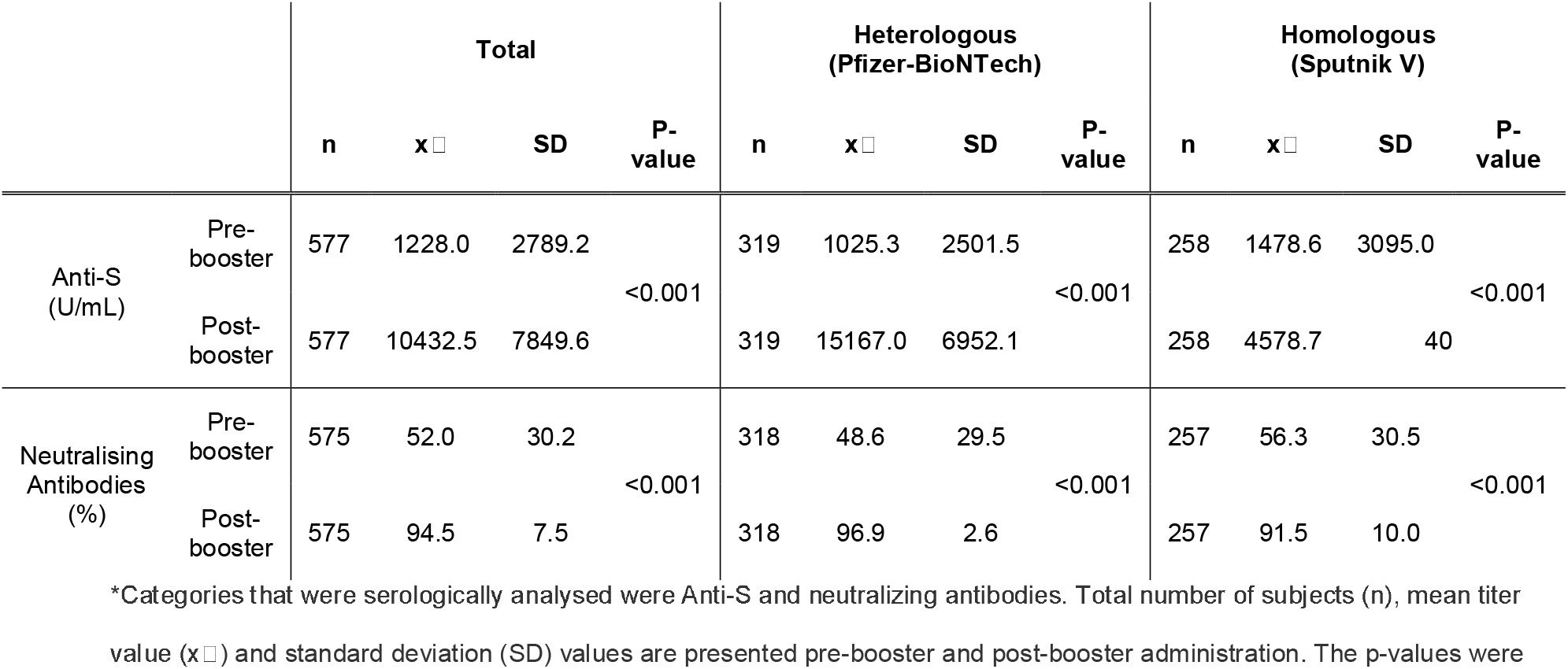
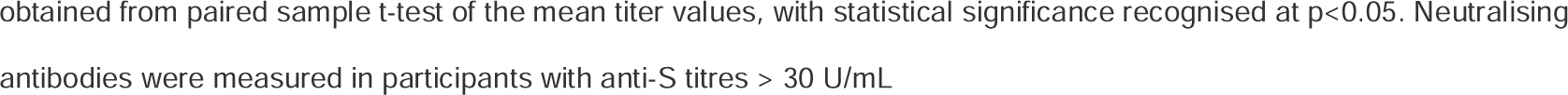
Comparison of serology results before and after booster administration for overall and by trial arm*. the homologous arm (p=0.002).

|  |  | Total |  |  |  | Heterologous<br>(Pfizer-BioNTech) |  |  |  | Homologous<br>(Sputnik V) |  |  |  |
| --- | --- | --- | --- | --- | --- | --- | --- | --- | --- | --- | --- | --- | --- |
| | | n | x$\bar{}$ | SD | P-value | n | x$\bar{}$ | SD | P-value | n | x$\bar{}$ | SD | P-value |
| Anti-S<br>(U/mL) | Pre-booster | 577 | 1228.0 | 2789.2 | <0.001 | 319 | 1025.3 | 2501.5 | <0.001 | 258 | 1478.6 | 3095.0 | <0.001 |
|  | Post-booster | 577 | 10432.5 | 7849.6 |  | 319 | 15167.0 | 6952.1 |  | 258 | 4578.7 | 40 |  |
| Neutralising<br>Antibodies<br>(%) | Pre-booster | 575 | 52.0 | 30.2 | <0.001 | 318 | 48.6 | 29.5 | <0.001 | 257 | 56.3 | 30.5 | <0.001 |
|  | Post-booster | 575 | 94.5 | 7.5 |  | 318 | 96.9 | 2.6 |  | 257 | 91.5 | 10.0 |  |

\*Categories that were serologically analysed were Anti-S and neutralizing antibodies. Total number of subjects (n), mean titer value ( $\bar{x}$ ) and standard deviation (SD) values are presented pre-booster and post-booster administration. The p-values were
obtained from paired sample t-test of the mean titer values, with statistical significance recognised at $p < 0.05$ . Neutralising antibodies were measured in participants with anti-S titres $> 30$ U/mL

## DISCUSSION

The results of this observational study show that both homologous and heterologous boosting are effective in eliciting an immunological response. Nonetheless, heterologous boosting with Pfizer-BioNTech in this cohort showed a higher immunological response, with more adverse effects. Through a search of the literature, no studies have been found for heterologous boosting for the Sputnik V vaccines, and hence compared our results to other heterologous boosting studies for SARS-CoV-2.

Firstly, from our assessment of reactogenicity, we observed overall low severity of adverse events in both trial arms, with local injection site being the most common adverse event. This result is comparable to adverse events documented in other reports comparing homologous and heterologous boosting (9, 13, 17-19), and each vaccine independently (20, 21). The incidence of adverse events was consistently higher in those aged <40 years, which is a common trend in SARS-CoV-2 vaccination studies (20, 22). More adverse events were reported in heterologous boosting compared to homologous boosting (p<0.001). This again was expected and is comparable to results from similar studies using Pfizer-BioNTech heterologous boosting in the context of ChAdOx1 nCoV-19 vaccination (9-11, 18, 23, 24). No adverse events were reported in participants aged 60+ in heterologous boosting, and one was recorded in participants aged 60+ in homologous boosting. However, the sample sizes, n=15 and n=11, respectively, were not sufficient to reliably comment on reactogenicity in this age group. Finally, we observed a peak in adverse events recorded on Day 28 of the trial, which does not compare to previous studies. Most studies constituted a time period of less than 28 days after boosting (11, 13, 19, 25) which may mean this finding has been missed due to limited study periods. A few reports have study periods of 28 days or longer, however, the reactogenicity was not reported per day of the study period (10, 18, 24), or not assessed as part of the study (12, 26). Nonetheless, it is important to consider that our finding may be confounded by the final appointment on day 28 being conducted face-to-face, since evidence shows that patients are more likely to report symptoms when face-to-face compared to call consults (27, 28). More heterologous boosting studies are required with longer time periods to establish the reliability of this finding.

Secondly, our immunogenicity analysis revealed increased Anti-S and neutralizing antibody titers in both arms of the trial. Of note, we observed significantly higher antibody titers in heterologous compared to homologous boosting. Although baseline titers for neutralizing antibodies were significantly higher in the homologous boosting cohort in our study, we still observed higher titers in the heterologous boosted cohort compared to the homologous boosted cohort, as observed in other studies (10, 11, 13, 15, 17, 23). Our results show homologous boosting in Sputnik V increased Anti-S titers by a factor of 3.1, and neutralizing antibody titers by a factor of 1.6. These values are similar to titers from other heterologous boosting studies, however, it is difficult to directly compare values as previous studies report titers in the context of other vaccines like AstraZeneca or mRNA1273 (Moderna). In contrast, these results are lower than titers expected for homologous vaccination using Sputnik V vaccine. (29, 30) This may have been affected by longer time periods between first dose and booster dose, as this was not be controlled for in this study. Furthermore, it is important to note that different detection kits, data presentation and processing methods were used in these studies compared to this. Evidence from homologous boosting with 0.3mL Pfizer-BioNTech dose shows an increase in protective neutralizing antibody by factor of 5.5 and 5 against the wildtype and delta SARS-CoV-2 variants (31). This shows that homologous boosting for Pfizer-BioNTech may be more immunologically advantageous, compared to using Pfizer-BioNTech as a heterologous booster or homologous boosting in Sputnik V vaccinated individuals. Nonetheless, it is important to note that all vaccination combinations demonstrate efficacy in eliciting immunological responses against SARS-CoV-2. Furthermore, data from Lapa D *et el*., shows that antibody titers are not statistically different between individuals fully vaccinated with Pfizer-BioNTech and those with Sputnik V (7). In consideration of all aforementioned evidence, in particular our results of increased immunogenicity with heterologous Sputnik V boosting, and the numerous reports of increased immunogenicity in heterologous boosting independent of type of vaccine used, we suggest that the variation in immunogenicity is more likely a consequence of heterologous boosting, as opposed to the specific type of booster vaccine used.

In summary, our results show that heterologous boosting of Sputnik V with Pfizer-BioNTech was both safe and efficacious. We observed a safe adverse reaction profile, with local injection site symptoms being the most commonly reported. This type of adverse event was reported at higher frequency amongst the heterologous boosted cohort, mostly occurring and peaking one day after receiving the injection. Heterologous boosting also yielded higher levels of antibodies than homologous boosting. These results can assist in guiding future selection of vaccine combinations, and are informative for individuals dual vaccinated with Sputnik V but yet to receive their booster dose. Nevertheless, expanding this study to investigate the inversed effect of Sputnik V boosting in Pfizer-BioNTech vaccination regimes, and other vaccination/booster combinations would be beneficial in guiding vaccine allocation, improving immunological defenses and overcoming SARS-CoV-2.

## Data Availability

All data produced in the present study are available upon reasonable request to the corresponding author

## LIMITATIONS AND FURTHER WORK

The main limitation of the trial is its non-blinded nature, incurring the potential risk of biased adverse events reporting, as participants’ knowledge of their vaccine may have influenced reactions. This was unlikely to have affected immunogenicity. However, we were unable to fully control the number of days from complete vaccination to booster administration, which may impact the antibody titers and hence immunogenicity analysis and make comparison to other studies less accurate. Furthermore, this study design allowed participants to choose their vaccines, resulting in significantly different demographics in each trial arm, which may have affected results. More studies are required to understand the observed effects of heterologous boosting, with a particular significance to research this in a wider variety of vaccines than currently available in the literature.

## Ethics approval

Approval granted after review from National COVID-19 Research and Ethics Committee (Approval Code: CRT-COVID2021-153).

## Consent to participate

Not applicable.

## Consent for publication

All authors approved the publishing of this data.

## Data availability

The datasets used and analysed in this study are available from the corresponding author on reasonable request. Requests may need prior approval from the ethical committee, and data may need to be de-identified.

## REFERENCES

1. World Health Organisation Director-General’s opening remarks at the media briefing on COVID-19 - 11 March 2020 [press release]. 2020.

2. WHO. World Health Organisation: Coronavirus Disease (COVID-19) Dashboard 2022 [12 March 2022]. Available from: https://covid19.who.int.

3. WHO. World Health Organisation: COVID-19 vaccine tracker and landscape. 2022.

4. Li Q, Nie J, Wu J, Zhang L, Ding R, Wang H, et al. SARS-CoV-2 501Y.V2 variants lack higher infectivity but do have immune escape. Cell. 2021;184(9):2362-71.e9.

5. Brown CM, Vostok J, Johnson H, Burns M, Gharpure R, Sami S, et al. Outbreak of SARS-CoV-2 Infections, Including COVID-19 Vaccine Breakthrough Infections, Associated with Large Public Gatherings - Barnstable County, Massachusetts, July 2021. MMWR Morb Mortal Wkly Rep. 2021;70(31):1059–62.

6. Kuhlmann C, Mayer CK, Claassen M, Maponga T, Burgers WA, Keeton R, et al. Breakthrough infections with SARS-CoV-2 omicron despite mRNA vaccine booster dose. Lancet. 2022;399(10325):625–6.

7. Lapa D, Grousova D, Matusali G, Meschi S, Colavita F, Bettini A, et al. Retention of Neutralizing response against SARS-CoV-2 Omicron variant in Sputnik V vaccinated individuals [PREPRINT]. 2022.

8. ECDC. European Centre for Disease Prevention and Control. Overview of EU/EEA country recommendations on COVID-19 vaccination with Vaxzevria, and a scoping review of evidence to guide decision-making. Stockholm: ECDC; 2021.

9. Nguyen TT, Quach THT, Tran TM, Phuoc HN, Nguyen HT, Vo TK, et al. Reactogenicity and immunogenicity of heterologous prime-boost immunization with COVID-19 vaccine. Biomed Pharmacother. 2022;147:112650.

10. Liu X, Shaw RH, Stuart ASV, Greenland M, Aley PK, Andrews NJ, et al. Safety and immunogenicity of heterologous versus homologous prime-boost schedules with an adenoviral vectored and mRNA COVID-19 vaccine (Com-COV): a single-blind, randomised, non-inferiority trial. Lancet. 2021;398(10303):856–69.

11. Benning L, Töllner M, Hidmark A, Schaier M, Nusshag C, Kälble F, et al. Heterologous ChAdOx1 nCoV-19/BNT162b2 Prime-Boost Vaccination Induces Strong Humoral Responses among Health Care Workers. Vaccines (Basel). 2021;9(8).

12. Vallée A, Vasse M, Mazaux L, Bonan B, Amiel C, Zia-Chahabi S, et al. An Immunogenicity Report for the Comparison between Heterologous and Homologous Prime-Boost Schedules with ChAdOx1-S and BNT162b2 Vaccines. J Clin Med. 2021;10(17).

13. Hillus D, Schwarz T, Tober-Lau P, Vanshylla K, Hastor H, Thibeault C, et al. Safety, reactogenicity, and immunogenicity of homologous and heterologous prime-boost immunisation with ChAdOx1 nCoV-19 and BNT162b2: a prospective cohort study. Lancet Respir Med. 2021;9(11):1255–65.

14. Barros-Martins J, Hammerschmidt SI, Cossmann A, Odak I, Stankov MV, Morillas Ramos G, et al. Immune responses against SARS-CoV-2 variants after heterologous and homologous ChAdOx1 nCoV-19/BNT162b2 vaccination. Nat Med. 2021;27(9):1525–9.

15. Dimeglio C, Herin F, Da-Silva I, Jougla I, Pradere C, Porcheron M, et al. Heterologous ChAdOx1-S/BNT162b2 Vaccination: Neutralizing Antibody Response to Severe Acute Respiratory Syndrome Coronavirus 2 (SARS-CoV-2). Clin Infect Dis. 2022;74(7):1315–6.

16. Faul F, Erdfelder E, Lang AG, Buchner A. G*Power 3: a flexible statistical power analysis program for the social, behavioral, and biomedical sciences. Behav Res Methods. 2007;39(2):175–91.

17. Atmar RL, Lyke KE, Deming ME, Jackson LA, Branche AR, El Sahly HM, et al. Homologous and Heterologous Covid-19 Booster Vaccinations. N Engl J Med. 2022.

18. Shaw RH, Stuart A, Greenland M, Liu X, Nguyen Van-Tam JS, Snape MD, et al. Heterologous prime-boost COVID-19 vaccination: initial reactogenicity data. Lancet. 2021;397(10289):2043–6.

19. Groß R, Zanoni M, Seidel A, Conzelmann C, Gilg A, Krnavek D, et al. Heterologous ChAdOx1 nCoV-19 and BNT162b2 prime-boost vaccination elicits potent neutralizing antibody responses and T cell reactivity against prevalent SARS-CoV-2 variants. EBioMedicine. 2022;75:103761.

20. Polack FP, Thomas SJ, Kitchin N, Absalon J, Gurtman A, Lockhart S, et al. Safety and Efficacy of the BNT162b2 mRNA Covid-19 Vaccine. N Engl J Med. 2020;383(27):2603–15.

21. Yegorov S, Kadyrove I, Negmetzhanov B, Kolesnikova Y, Kolescickenko S, Korchukov I, et al. Sputnik-V reactogenicity and immunogenicity in the blood and mucosa: a prospective cohort study. [Pre-print]2022.

22. Beatty AL, Peyser ND, Butcher XE, Cocohoba JM, Lin F, Olgin JE, et al. Analysis of COVID-19 Vaccine Type and Adverse Effects Following Vaccination. JAMA Netw Open. 2021;4(12):e2140364.

23. Nordström P, Ballin M, Nordström A. Effectiveness of heterologous ChAdOx1 nCoV-19 and mRNA prime-boost vaccination against symptomatic Covid-19 infection in Sweden: A nationwide cohort study. Lancet Reg Health Eur. 2021;11:100249.

24. Li J, Hou L, Guo X, Jin P, Wu S, Zhu J, et al. Heterologous AD5-nCOV plus CoronaVac versus homologous CoronaVac vaccination: a randomized phase 4 trial. Nat Med. 2022;28(2):401–9.

25. Schmidt T, Klemis V, Schub D, Mihm J, Hielscher F, Marx S, et al. Immunogenicity and reactogenicity of heterologous ChAdOx1 nCoV-19/mRNA vaccination. Nat Med. 2021;27(9):1530–5.

26. Yorsaeng R, Vichaiwattana P, Klinfueng S, Wongsrisang L, Sudhinaraset N, Vongpunsawad S, et al. Immune response elicited from heterologous SARS-CoV-2 vaccination: Sinovac (CoronaVac) followed by AstraZeneca (Vaxzevria). medRxiv 20212021.

27. McKinstry B, Hammersley V, Burton C, Pinnock H, Elton R, Dowell J, et al. The quality, safety and content of telephone and face-to-face consultations: a comparative study. Qual Saf Health Care. 2010;19(4):298–303.

28. Leng S, MacDougall M, McKinstry B. The acceptability to patients of video-consulting in general practice: semi-structured interviews in three diverse general practices. J Innov Health Inform. 2016;23(2):141.

29. Tukhvatulin AI, Dolzhikova IV, Shcheblyakov DV, Zubkova OV, Dzharullaeva AS, Kovyrshina AV, et al. An open, non-randomised, phase 1/2 trial on the safety, tolerability, and immunogenicity of single-dose vaccine “Sputnik Light” for prevention of coronavirus infection in healthy adults. Lancet Reg Health Eur. 2021;11:100241.

30. Komissarov AA, Dolzhikova IV, Efimov GA, Logunov DY, Mityaeva O, Molodtsov IA, et al. Boosting of the SARS-CoV-2-Specific Immune Response after Vaccination with Single-Dose Sputnik Light Vaccine. J Immunol. 2022;208(5):1139–45.

31. Falsey AR, Frenck RW, Walsh EE, Kitchin N, Absalon J, Gurtman A, et al. SARS-CoV-2 Neutralization with BNT162b2 Vaccine Dose 3. N Engl J Med. 2021;385(17):1627–9.

